# The Effect of a U.S. Poverty Reduction Intervention on Maternal Assessments of Young Children’s Health, Nutrition, and Sleep: A Randomized Control Trial

**DOI:** 10.1101/2023.05.25.23290530

**Authors:** Jessica F. Sperber, Lisa A. Gennetian, Emma R. Hart, Alicia Kunin-Batson, Katherine Magnuson, Greg J. Duncan, Hirokazu Yoshikawa, Nathan A. Fox, Sarah Halpern-Meekin, Kimberly G. Noble

## Abstract

**Importance:** Children experiencing poverty are more likely to experience worse health outcomes during the first few years of life, including injury, chronic illness, worse nutrition, and poorer sleep. The extent to which a poverty reduction intervention improves children’s health, nutrition, sleep, and healthcare utilization is unknown.

**Objective:** To determine the effect of a 3-year, monthly unconditional cash transfer on health, nutrition, sleep, and healthcare utilization of children experiencing poverty who are healthy at birth.

**Design:** Longitudinal randomized control trial

**Setting:** Mother-infant dyads were recruited from postpartum wards in 12 hospitals in four cities across the U.S.

**Participants:** 1,000 mothers were enrolled in the study. Eligibility criteria included: an annual income below the federal poverty line, being of legal age for consent, speaking English or Spanish, residing in the state of recruitment, and having an infant admitted to the well-baby nursery with plans to be discharged to the custody of the mother.

**Intervention:** Mothers were randomly assigned to receive either a high-cash gift ($333 per month, or $3,996 per year; *n*=400) or a low-cash gift ($20 per month, or $240 per year; *n*=600) for the first several years of their child’s life.

**Main Outcomes and Measures:** Pre-registered maternal assessments of the focal child’s health, nutrition, sleep, and healthcare utilization were collected at children’s ages 1, 2, and 3.

**Results:** Enrolled participants were majority Black (42%) and Hispanic (41%). 857 mothers participated in all three waves of data collection. We found no statistically detectable differences between the high-cash and low-cash gift groups in maternal assessments of children’s overall health, sleep, or healthcare utilization. However, mothers in the high-cash gift group reported higher child consumption of fresh produce compared with mothers in the low-cash gift group at age 2, the only time point it was measured (*β=*0.17, SE=0.07, *p*=0.03).

**Conclusions and Relevance:** In this RCT, unconditional cash transfers to mothers experiencing poverty did not improve their reports of their child’s health, sleep, or healthcare utilization. However, stable income support of this magnitude improved toddler’s consumption of fresh produce. Healthy newborns tend to grow into healthy toddlers, and the impacts of poverty reduction on children’s health and sleep may not be fully borne out until later in life.

**Trial Registration:** Baby’s First Years (BFY; ID NCT03593356) https://clinicaltrials.gov/ct2/show/NCT03593356?term=NCT03593356&draw=2&rank=1

**Key points:** *Question:* Does poverty reduction improve health, nutrition, and sleep in young children?

*Findings:* In this RCT of 1,000 mother-child dyads experiencing poverty, a monthly unconditional cash transfer did not improve children’s health or sleep in the first three years of life. However, the cash transfers led to increased consumption of fresh produce.

*Meaning:* Among children experiencing poverty, a monthly cash gift affected healthy food intake, but not health or sleep. Most children had few health problems, though emergent medical care use was high.

## Introduction

Children experiencing poverty are more likely to experience worse health outcomes, including injury,^1^ chronic illness,^2^ and poor sleep^3^ and are more likely to use emergency health services.^4^ Numerous factors likely contribute to the associations between poverty and children’s health, including poor prenatal care,^5^ exposure to environmental toxicants,^6^ low quality housing and neighborhoods,^7^ and lack of access to medical care^7^ and nutritious foods.^8^ Poverty may also impact young children’s sleep through parental mental health^9^ and the quality of the sleep environment^10^ and bedtime routines.^9,10^

Previous quasi-experimental work suggests even small increases in income may improve health trajectories for children experiencing poverty. For example, expansion of the earned income tax credit (EITC), the largest anti-poverty policy in the U.S. prior to pandemic-related tax expansions, reduced the incidence of low-birthweight infants in the population.^11^ Similarly, exogenous increases in minimum wage were found to improve children’s nutritional status in low-income countries.^12^ However, the health impacts of poverty reduction for otherwise healthy infants in the U.S. is unknown.

Monetary investment and improvements in parental stress are two potential pathways by which we have theorized that poverty reduction may improve developmental trajectories.^13^ For example, greater financial resources may allow parents to invest in high-quality inputs that support child health and sleep, including nutritious foods, preventive medical care, a separate sleep space, and safe housing. Additionally, reduced financial strain may reduce parental stress and mental health symptoms, which may subsequently improve the quality of family interactions, bedtime routines, and child sleep quality.

The present study evaluates the effect of a poverty reduction intervention on children’s health, nutrition, sleep, and healthcare utilization in the first three years of life. Such an intervention exemplifies a scalable public health approach. We hypothesized that a monthly, unconditional cash transfer would improve maternal assessments of children’s health, nutrition, and sleep, and reduce use of emergency healthcare.

## Methods

### Study Design

Baby’s First Years (BFY) is a parallel-group, randomized control trial of poverty reduction. Between May 2018 and June 2019, 1,000 mothers experiencing poverty were recruited from postpartum wards after giving birth, and were offered a monthly unconditional cash transfer (referred to as a “cash gift”). Mothers were randomly assigned to either a high-cash gift group (*n*=400) or a low-cash gift group (*n*=600). The monthly gifts were initially promised for the first 40 months of their child’s life, and were subsequently extended through 76 months. More information on study design can be found in eMethods in the Supplement and in Noble et al.^13^

Upon providing written informed consent, mothers completed a baseline interview and received compensation for their participation ($50). For the next three years, mothers were invited to complete annual surveys around the time of their child’s birthday. All study procedures were approved by the Institutional Review Boards of [blinded for review].

### Participants

Mothers were recruited from 12 hospitals across 4 U.S. metropolitan areas: New York, Omaha, New Orleans, and Minneapolis/St. Paul. Eligibility criteria included: 1) being of legal age to provide informed consent; 2) reporting a household income below the federal poverty line during the previous year; 3) able to speak English or Spanish; 4) the infant not being admitted to the neonatal intensive care unit; 5) residing in the state of recruitment; 6) planning to remain in-state within the next year, and 7) infant to be discharged into the mother’s custody.

### Randomization and Masking

Randomization occurred within hospitals, with 60% randomized to the low-cash gift group and 40% randomized to the high-cash gift group. Interviewers retrieved assignment after obtaining informed consent. Interviewers could not be masked to condition during recruitment; however, they were not informed or reminded of participants’ treatment status during follow-up assessments.

### Intervention

Mothers in the high-cash gift group received $333 per month ($3,996 per year), whereas mothers in the low-cash gift group received $20 per month ($240 per year). Funds were disbursed monthly onto an electronic debit card (branded “4MyBaby”).^15^ The receipt of $4,000 in cash gifts increased the average family’s income by ∼20%.^16^ Provisions instituted by state agencies and legislation ensured the cash gifts affected neither mothers’ eligibility for or amount of public benefits.

Participants were informed they could spend the money how they wished, and receipt of the funds was not conditioned upon continued participation in the study. Among those who consented to allow access to transaction data (*n*=900), only five families (all in the low-cash gift group) have not withdrawn funds from the debit card three years post-randomization.

### Measures

Each year, mothers completed surveys of their child’s health, nutrition, sleep, and healthcare utilization. Such parent-reported measures are frequently used as proxies for child health outcomes (e.g., Page et al.^17^) and correlate with objective assessments.^17,18^ The primary outcomes of interest were the measures of health, nutrition, sleep, and healthcare utilization at each wave described below. The secondary outcome of interest was a Poor Health Index (described in the Supplement). These outcomes were preregistered at clinicaltrials.gov (ID: NCT03593356).

Four global measures of health, nutrition, sleep, and healthcare utilization were also used, though these composite measures were not pre-registered. Each composite was formed by standardizing the outcomes in that domain, averaging them together, and then re-standardizing. Standardization was performed separately at each wave of data collection, across both high-cash and low-cash gift groups. Higher values indicate poorer outcomes in that domain.

#### Health Outcomes – Ages 1, 2, and 3

##### Overall Health

Mothers rated their child’s overall health on a 5-point Likert scale, from 1 (*excellent*) to 5 (*poor*).

##### Diagnosis with a health condition or disability

Mothers indicated via a dichotomous item whether their child was diagnosed with a health condition or disability since birth. Mothers who endorsed “yes” on the item were asked to specify the diagnosis, and these responses were coded.

#### Nutrition Outcomes – Age 2

On the Age-2 survey, mothers responded to 4-items regarding their child’s food intake. The items were adapted from the Los Angeles County WIC Survey.^18^

##### Healthy Foods

Two items assessed how frequently their child ate fruits and vegetables on an average day. Frequency was assessed via a Likert scale ranging from 0 (*never*) to 5 (*5+ times per day*). We created an additive index by summing the total number of times the child consumed produce each day.

##### Unhealthy Foods

Two items assessed how frequently their child consumed sweets (e.g., sweetened cereals, fruit bars, etc.) and sweetened drinks (e.g., juice, chocolate milk) on an average day. Frequency was assessed via a Likert scale ranging from 0 (*never*) to 5 (*5+ times per day*). We created an additive index by summing the total number of times the child consumed unhealthy foods each day.

#### Sleep Outcomes – Ages 1, 2, and 3

##### Sleep Disturbances

Sleep disturbances were assessed through the PROMIS Sleep Disturbance-Short Form,^19^ consisting of four items assessing the frequency of sleep-related difficulties over the last 7 days using a 5-point Likert scale, 1 (*never*) to 5 (*always*). One positively-stated item was reverse coded before being summed. Higher scores indicated more sleep disturbances. Mothers needed to respond to at least three of the four items to obtain a valid score. Due to an administrative error, one item was excluded from the Age-3 survey. Thus, the score at Age 3 reflects the sum of only 3 items, rather than 4.

##### Poor Bedtime Routines

Bedtime routines were indexed via two items from the Confusion, Hubbub, and Order Scale administered at Age 1 and Age 2 (CHAOS).^21^ Participants selected whether the statements “We have an evening bedtime routine” and “My child goes to bed at a regular time” were true (or false) of their home most of the time. We averaged the scores of these two items together, with higher scores indicating fewer sleep routines.

#### Healthcare Utilization Outcomes – Ages 1, 2, and 3

##### Doctor Visits

Mothers responded to two items indexing the number of times they brought their child to a doctor due to injury or illness in the past year (*0–1 visits, 2–5 visits*, or *6+ visits*). Across all three waves of data collection, most mothers (92%+) reported less than 6+ doctor visits in the last year. We collapsed these items into two dichotomous indicators, representing whether the mother reported 2+ doctor visits due to illness or injury in the past year.

##### Emergency Room/Urgent Care Visits

Mothers reported the number of times they brought their child to an emergency room (ER) or urgent care center in the past year using a categorical indicator (*0 visits, 1 visit, 2–5 visits*, or *6+ visits*). Across waves of data collection, between 35–54% of mothers reported at least one visit, and nearly all mothers (98–99%) reported fewer than 6+ visits. For analyses, we created an ordinal variable reflecting whether the mother reported 0, 1, or 2+ ER/urgent care visits in the last year.

### Missing Data

High response rates were observed at annual follow-up assessments: after adjusting for mother-child separations (*n*=2), maternal incarcerations (*n*=4), and infant deaths (*n*=4), at least 92% of the sample participated in each timepoint (see CONSORT diagrams for follow-up waves in eFigure 1 in the Supplement). 857 mothers completed all three surveys.

### Statistical Analysis

We use two-tailed OLS regression equations with robust standard errors to estimate the effects of the cash transfer on individual and composite outcomes at ages 1, 2, and 3. Additionally, we analyzed cumulative impacts of the intervention by pooling across waves for each outcome (i.e., ages 1, 2, and 3; *n*=2,768). Analysis of the preregistered Poor Health Index, as well exploratory analyses estimating the impact of the cash gift on post-birth diagnoses, childhood vaccinations, missed medical/dental care, Medicaid receipt, and children’s consumption of cow’s milk, are in the Supplement.

Intent-to-treat (ITT) analyses were conducted. Effect sizes reflecting the standardized treatment impact, divided by the standard deviation (SD) of the control group, are reported in Table 3. Marginal effects derived from probit regressions of dichotomous outcomes are presented in eTable 1 in the Supplement. All analyses were adjusted for 27 preregistered covariates measured at baseline (see Table 1). We also adjust for the child’s exact age at the time of the interview. The COVID-19 pandemic began partway through in-person Age-1 data collection, at which point data collection pivoted from in-person to phone surveys (all Age-2 and Age-3 surveys were collected remotely). We include a dummy variable for survey administration method in the Age-1 wave.

**Table 1.** Descriptive Statistics at Baseline^a^

|  | **Low-Cash Gift Group** |  | **High-Cash Gift Group** |  |
| --- | --- | --- | --- | --- |
|  | *Mean (SD)* | *N* | *Mean (SD)* | *N* |
| Child is female | 0.50 | 600 | 0.48 | 400 |
| Child weight at birth (pounds) | 7.10 (1.08) | 599 | 7.10 (1.01) | 399 |
| Child gestational age (weeks) | 39.10 (1.25) | 596 | 39.00 (1.24) | 399 |
| Mother age at birth (years) | 26.80 (5.82) | 600 | 27.40 (5.87) | 400 |
| Mother education (years) | 11.90 (2.83) | 593 | 11.90 (2.96) | 398 |
| Mother race/ethnicity: white, non-Hispanic | 0.11 | 600 | 0.09 | 400 |
| Mother race/ethnicity: Black, non-Hispanic | 0.40 | 600 | 0.44 | 400 |
| Mother race/ethnicity: multiple, non-Hispanic | 0.04 | 600 | 0.03 | 400 |
| Mother race/ethnicity: other or unknown | 0.05 | 600 | 0.03 | 400 |
| Mother race/ethnicity: Hispanic | 0.41 | 600 | 0.42 | 400 |
| Mother marital status: never married | 0.43 | 600 | 0.50 | 400 |
| Mother marital status: single, living with partner | 0.26 | 600 | 0.22 | 400 |
| Mother marital status: married | 0.21 | 600 | 0.22 | 400 |
| Mother marital status: divorced/separated | 0.05 | 600 | 0.03 | 400 |
| Mother marital status: Other family structure or status unknown | 0.06 | 600 | 0.05 | 400 |
| Mother health is good or better | 0.88 | 600 | 0.92 | 400 |
| Mother depression (CESD) | 6.80 (4.52) | 600 | 6.90 (4.61) | 400 |
| Cigarettes per week during pregnancy | 5.00 (21.17) | 595 | 3.50 (11.76) | 397 |
| Alcohol drinks per week during pregnancy | 0.20 (1.63) | 598 | 0.00 (0.39) | 399 |
| Number of children born to mother | 2.40 (1.38) | 600 | 2.50 (1.41) | 400 |
| Number of adults in household | 2.10 (1.00) | 600 | 2.00 (0.96) | 400 |
| Biological father lives in household | 0.38 | 600 | 0.35 | 400 |
| Household combined income | 22,466 (21,360) | 562 | 20,918 (16,146) | 370 |
| Household income unknown | 0.06 | 600 | 0.08 | 400 |
| Household net worth | -1,981 (28,640) | 531 | -3,308 (20,323) | 358 |
| Household net worth unknown | 0.12 | 600 | 0.11 | 400 |

Notes: <sup>a</sup> Table adapted from Noble et al.<sup>13</sup>

**Table 2.** Descriptive Statistics of Outcome Variables at Ages 1, 2, and 3 by Treatment Status

|  | **Age 1** |  | **Age 2** |  | **Age 3** |  |
| --- | --- | --- | --- | --- | --- | --- |
|  | *Low-Cash*<br>*Gift Group* | *High-Cash*<br>*Gift Group* | *Low-Cash*<br>*Gift Group* | *High-Cash*<br>*Gift Group* | *Low-Cash*<br>*Gift Group* | *High-Cash*<br>*Gift Group* |
| ***Child Health*** |  |  |  |  |  |  |
| Maternal rating of child's overall health (1=excellent, 5=poor) | 1.49<br>(0.73) | 1.51<br>(0.80) | 1.53<br>(0.76) | 1.51<br>(0.80) | 1.56<br>(0.82) | 1.61<br>(0.87) |
| Maternal report of whether child has a diagnosis of health condition or disability (%) | 0.12 | 0.15 | 0.06 | 0.07 | 0.10 | 0.11 |
| Overall Poor Health Index | 2.93<br>(1.54) | 3.10<br>(1.72) | 2.48<br>(1.48) | 2.48<br>(1.54) | 2.51<br>(1.65) | 2.53<br>(1.55) |
| ***Nutrition<sup>a</sup>*** |  |  |  |  |  |  |
| Healthy foods (Number of times consumed per day) | N/A | N/A | 4.18<br>(2.12) | 4.50<br>(2.16) | N/A | N/A |
| Unhealthy foods (Number of times consumed per day) | N/A | N/A | 3.45<br>(2.21) | 3.48<br>(2.15) | N/A | N/A |
| ***Sleep*** |  |  |  |  |  |  |
| PROMIS-Sleep Disturbance Scale<sup>b</sup> | 8.02<br>(3.44) | 7.70<br>(3.36) | 7.49<br>(3.47) | 7.61<br>(3.35) | 5.04<br>(2.61) | 5.10<br>(2.50) |
| Bedtime Routines<sup>c</sup> | 0.16<br>(0.30) | 0.15<br>(0.29) | 0.16<br>(0.31) | 0.16<br>(0.31) | N/A | N/A |
| ***Healthcare Utilization*** |  |  |  |  |  |  |
| 2+ doctor visits due to illness per year (%) | 0.52 | 0.56 | 0.31 | 0.31 | 0.32 | 0.30 |
| 2+ doctor visits due to injury per year (%) | 0.02 | 0.02 | 0.03 | 0.03 | 0.04 | 0.02 |
| ER/Urgent Care Visits per year | 0.78<br>(0.83) | 0.86<br>(0.85) | 0.54<br>(0.74) | 0.56<br>(0.76) | 0.49<br>(0.74) | 0.50<br>(0.73) |
| **Sample Size** | **547** | **382** | **543** | **376** | **542** | **378** |

*Note:* Mean (SD). Except for healthy food consumption, larger values indicate poorer outcomes in each domain.
<sup>a</sup> Nutrition items were only administered on the Age-2 survey.
<sup>b</sup> One item was mistakenly excluded from the PROMIS-Sleep Disturbance Scale on the Age-3 survey, resulting in a lower average score at that age.
<sup>c</sup> Bedtime routines represents the average of two items from the CHAOS questionnaire, which was only administered at the Age-1 and Age-2 surveys.

**Table 3.**
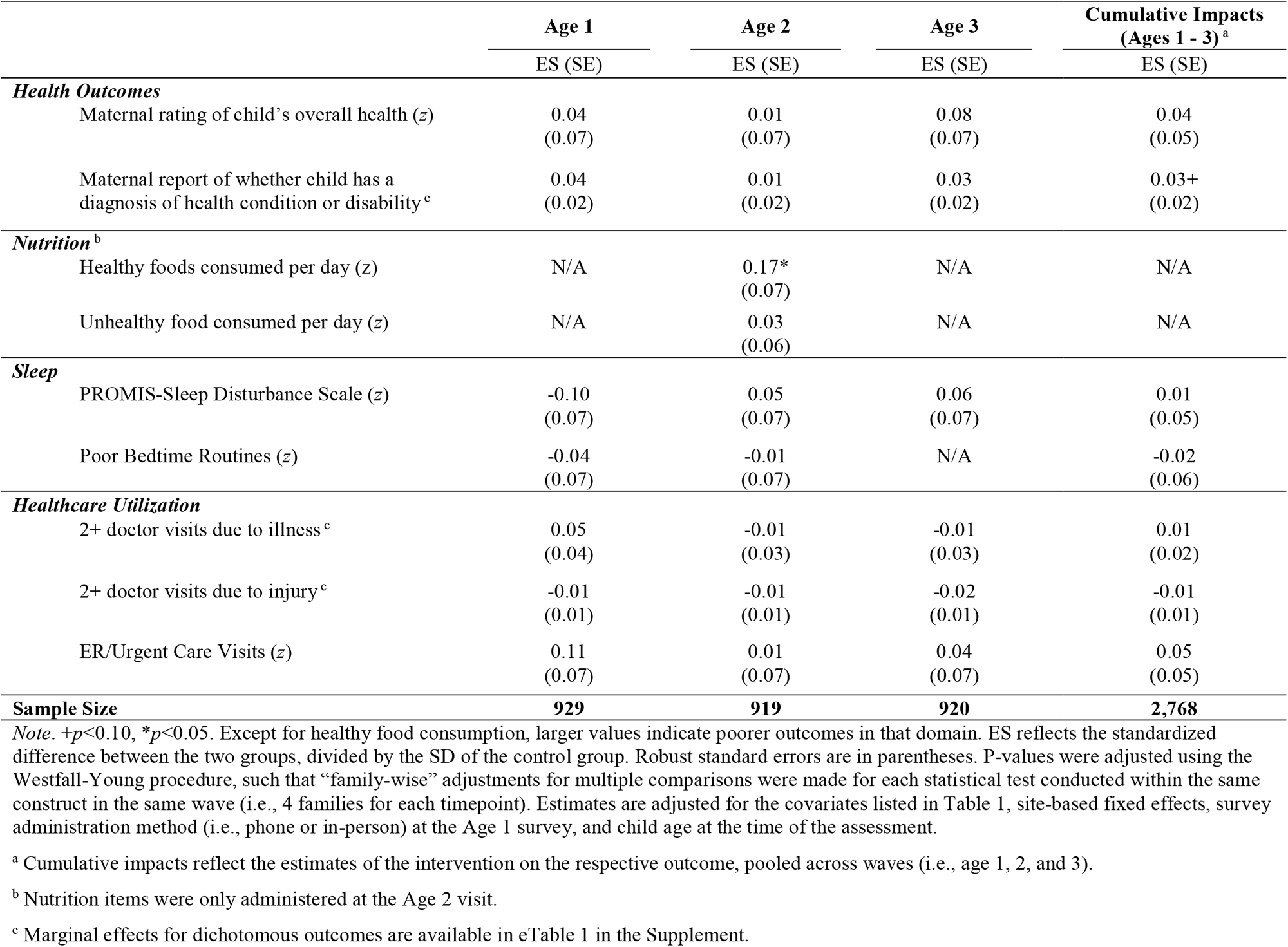
Intent-to-Treat (ITT) Impacts of Unconditional Cash Transfer on Child Health, Nutrition, Sleep, and Healthcare Utilization

|  | Age 1 | Age 2 | Age 3 | Cumulative Impacts<br>(Ages 1 - 3)<sup>a</sup> |
| --- | --- | --- | --- | --- |
|  | ES (SE) | ES (SE) | ES (SE) | ES (SE) |
| **Health Outcomes** |  |  |  |  |
| Maternal rating of child's overall health (z) | 0.04<br>(0.07) | 0.01<br>(0.07) | 0.08<br>(0.07) | 0.04<br>(0.05) |
| Maternal report of whether child has a diagnosis of health condition or disability<sup>c</sup> | 0.04<br>(0.02) | 0.01<br>(0.02) | 0.03<br>(0.02) | 0.03+<br>(0.02) |
| **Nutrition<sup>b</sup>** |  |  |  |  |
| Healthy foods consumed per day (z) | N/A | 0.17*<br>(0.07) | N/A | N/A |
| Unhealthy food consumed per day (z) | N/A | 0.03<br>(0.06) | N/A | N/A |
| **Sleep** |  |  |  |  |
| PROMIS-Sleep Disturbance Scale (z) | -0.10<br>(0.07) | 0.05<br>(0.07) | 0.06<br>(0.07) | 0.01<br>(0.05) |
| Poor Bedtime Routines (z) | -0.04<br>(0.07) | -0.01<br>(0.07) | N/A | -0.02<br>(0.06) |
| **Healthcare Utilization** |  |  |  |  |
| 2+ doctor visits due to illness<sup>c</sup> | 0.05<br>(0.04) | -0.01<br>(0.03) | -0.01<br>(0.03) | 0.01<br>(0.02) |
| 2+ doctor visits due to injury<sup>c</sup> | -0.01<br>(0.01) | -0.01<br>(0.01) | -0.02<br>(0.01) | -0.01<br>(0.01) |
| ER/Urgent Care Visits (z) | 0.11<br>(0.07) | 0.01<br>(0.07) | 0.04<br>(0.07) | 0.05<br>(0.05) |
| **Sample Size** | **929** | **919** | **920** | **2,768** |

*Note.* + $p < 0.10$ , \* $p < 0.05$ . Except for healthy food consumption, larger values indicate poorer outcomes in that domain. ES reflects the standardized difference between the two groups, divided by the SD of the control group. Robust standard errors are in parentheses. P-values were adjusted using the Westfall-Young procedure, such that “family-wise” adjustments for multiple comparisons were made for each statistical test conducted within the same construct in the same wave (i.e., 4 families for each timepoint). Estimates are adjusted for the covariates listed in Table 1, site-based fixed effects, survey administration method (i.e., phone or in-person) at the Age 1 survey, and child age at the time of the assessment.
<sup>a</sup> Cumulative impacts reflect the estimates of the intervention on the respective outcome, pooled across waves (i.e., age 1, 2, and 3).
<sup>b</sup> Nutrition items were only administered at the Age 2 visit.
<sup>c</sup> Marginal effects for dichotomous outcomes are available in eTable 1 in the Supplement.

Adjustments for multiple comparisons were made using the Westfall-Young^20^ procedure when analyzing measures within the same construct (i.e., health, nutrition, sleep, or healthcare utilization) in the same wave. Adjusted p-values are featured in the results below. Data are publicly available at the Inter-university Consortium for Political and Social Research (ICPSR; ID 37871). Analyses were conducted using Stata.^21^

## Results

Recruitment occurred between May 2018 and June 2019. Details are in the previously published baseline consolidated standards of reporting trials (CONSORT) diagram.^13^

### Descriptives

Of the 1,000 randomized mothers, 1.2% self-identified as American Indian/ Eskimo/Aleut, 0.9% as Asian or Pacific Islander, 41.5% as Black, 10.1% as White, 3.6% as being multiple races, and 1.7% as being some other race. Of Hispanic-identifying participants (40.9%), most identified as from either the Dominican Republic (34%), the United States (33%), or Mexico (16%). 49.2% of the infants were female.

Table 1 presents descriptive statistics by treatment status for all participants at baseline (*n*=1,000). On average, infants were of normal birth weight (*M*=7 pounds, *SD*=1.10) and born at term (*M*=39 weeks, *SD*=1.30 weeks). Four mothers reported their child as deceased by the Age-1 visit (three infants in the high-cash gift group). This mortality rate is notably higher than the national average of 2.03 deaths for every 1,000 full-term infants.^22^

Descriptive statistics for the main outcomes are presented in Table 2. On average, 63% of mothers rated their child’s overall health as excellent, 23% rated it as very good, and 11% rated it as good. Across all visits, 2–4% of mothers rated their child’s health as fair or poor, which aligns with the national average of 2.2% (95% CI 1.7, 2.8).^23^ By Age 3, 10% of mothers reported that their child had been diagnosed with a health condition or disability. Though national estimates vary widely, between 5–14% of all children have a disability, with those experiencing poverty demonstrating higher rates of diagnosis than more economically advantaged peers.^24^ Autism (*n*=28) and asthma (*n*=20) were the most frequently reported diagnoses by Age 3.

Across waves of data collection, between 15–28% of mothers reported bringing their child to an ER or urgent care center 2+ times in the past year. This is higher than national rates for children living in poverty nationwide: 11% of children living in poverty reported 2+ ER visits in 2019 (95% CI 9.2, 13.5), and 14.2% reported 2+ urgent care visits that same year (95% CI 11.8, 16.9).^23^

### Impacts on Health, Nutrition, Sleep, and Healthcare Utilization

The cash gift did not impact any child health, sleep, or healthcare utilization outcome at Ages 1, 2, or 3 (Table 3). However, receipt of the cash gift caused significantly higher child consumption of fruits and vegetables at Age 2, the only age at which it was measured, *β=*0.17, SE=0.07, *p*=0.03. Exploratory analyses revealed this effect was driven by increased fruit consumption (*β=*0.22, SE=0.07, *p*<0.01) rather than vegetable consumption (*β=*0.06, SE=0.07, *p*=0.41). The cash gift did not alter consumption of sweets and sweetened beverages, *β=*0.03, SE=0.06, *p*=0.69. No differences were observed by treatment status on any global measure of health, nutrition, sleep, or healthcare utilization at any timepoint (Table 4).

**Table 4.** Intent-to-Treat (ITT) Impacts of Unconditional Cash Transfer on Global Measures of Child Health, Nutrition, Sleep, and Healthcare Utilization

|  | **Age 1** | **Age 2** | **Age 3** | **Cumulative Impacts**<br>**(Age 1 – 3)<sup>a</sup>** |
| --- | --- | --- | --- | --- |
|  | ES (SE) | ES (SE) | ES (SE) | ES (SE) |
| Global Health<sup>b</sup> (z) | 0.10<br>(0.07) | 0.02<br>(0.07) | 0.11<br>(0.07) | 0.08<br>(0.05) |
| Global Nutrition<sup>c</sup> (z) | N/A | -0.10<br>(0.07) | N/A | N/A |
| Global Sleep<sup>d</sup> (z) | -0.10<br>(0.07) | 0.03<br>(0.07) | N/A | -0.04<br>(0.06) |
| Global Healthcare Utilization<sup>e</sup> (z) | 0.10<br>(0.07) | -0.01<br>(0.07) | -0.04<br>(0.07) | 0.01<br>(0.05) |
| **Sample Size** | **929** | **919** | **920** | **2,768** |

*Note.* + $p < 0.10$ . Each composite was formed by averaging together the relevant indicators and standardizing the mean, with higher values reflecting poorer outcomes in that domain. ES reflects the standardized difference between the two groups, divided by the SD of the control group. Robust standard errors are in parentheses. Estimates are adjusted for the covariates listed in Table 1, site-based fixed effects, survey administration method (i.e., phone or in-person) at the Age 1 survey, and child age at the time of the assessment.
<sup>a</sup> Cumulative Impacts reflect the estimates of the intervention on the respective outcome, pooled across waves (i.e., ages 1, 2, and 3).
<sup>b</sup> Global health consists of the overall health rating and the indicator of disability diagnosis.
<sup>c</sup> Global nutrition consists of the servings of unhealthy and healthy (reverse coded) foods consumed each day.
<sup>d</sup> Global sleep consists of the PROMIS-Sleep Disturbance Scale and two items relating to bedtime routines and consistent bedtimes from the CHAOS scale. The CHAOS scale was administered only at the Age 1 and Age 2 assessment; therefore, the only available Age 3 sleep measure was the PROMIS-Sleep Disturbance scale, and results for this are presented in Table 3.
<sup>e</sup> Global healthcare utilization consists of the 2+ doctor visits due to illness or injury indicators and the number of ER/urgent care visits.

## Discussion

This preregistered study found that three years of monthly, unconditional cash transfers for families experiencing poverty did not improve maternal reports of children’s health, sleep, or healthcare utilization. However, the cash gifts did lead to increased reported produce consumption at age 2. Previous work finds low-income mothers tend to perceive fresh produce as expensive and inaccessible,^25,26^ and may be averse to introducing novel foods that their children may reject.^27^ The cash gifts may have encouraged low-income mothers to take the financial risk of investing in fresh produce.

By design, this sample consisted of infants who did not require neonatal intensive care at birth, of whom the vast majority were born at-term, and of normal birthweight. It is therefore unsurprising that most mothers rated their children as having excellent or very good health, with rates of health conditions or disabilities aligned with national averages. Most studies linking poverty reduction to children’s health do not utilize such exclusionary criteria (e.g., Strully et al.^12^). It’s possible that positive impacts of the cash transfers on child health were less likely among this sample of children deemed healthy at birth.

Rates of emergency medical use among our sample were higher than the national average for low-income mothers. Structural barriers might interfere with preventive care use,^28^ leading to higher emergency health services utilization. A modest increase in monthly income may be insufficient to overcome the lack of a consistent medical home. Direct interventions that connect families with services may be more effective at reducing emergency healthcare utilization among children experiencing poverty (e.g., Goodman et al.^29^).

We have previously reported no effect of the cash gift on some potential pathways that may have improved outcomes these domains. For example, the cash gift did not improve maternal stress, maternal mental health symptoms, or maternal relationship quality,^30^ which are closely associated with children’s sleep quality and the quality of bedtime interactions.^9^ Although parents reported greater investment in child-focused expenditures (e.g., books and toys),^16^ the cash gift had no effect on reported food insecurity, housing quality, or the likelihood of purchasing a crib.^16^ The null effects observed on these hypothesized mechanisms may account for the null results observed for early childhood health, healthcare utilization, and sleep.

The effects of a poverty reduction intervention on children’s health may not emerge until later in development. Indeed, many illnesses associated with childhood poverty such as hypertension, type II diabetes, and heart disease tend to emerge in adolescence or adulthood.^31,32^ Natural experiments such as the Great Smokey Mountain Study have found that cash transfers during middle childhood are associated with positive effects on physical and mental health in adulthood.^33^ It will be important to follow the BFY sample to measure the extent to which investments in early childhood may reduce the incidence of disease processes that emerge later in life.^34,35^

### Strengths & Limitations

An important strength of this study is its large sample, experimental design, low rates of attrition, and preregistered analysis, which represents an improvement over prior studies that have examined associations between income and children’s health, sleep, and nutrition in cross-sectional or observational studies. A limitation is that it relies entirely on maternal assessments, which may introduce bias and be less reliable than objective assessments. However, parental report of children’s overall health, medical history, and sleep duration tends to be highly correlated with objective assessments, including abstraction of medical records and actigraphy.^36,37^ Another limitation is reflected in the limited variability on some survey items, potentially indicating that the response options provided were not ideally suited for assessing medical care in young children. We intend to follow the families through middle childhood and will continue to assess the children’s health and development over time.

## Data Availability

As of this writing, baseline, Age-1, and Age-2 data are publicly available at the Inter-university Consortium for Political and Social Research (ICPSR) repository (ID 37871)

https://doi.org/10.3886/ICPSR37871.v4

## Acknowledgements

We thank the University of Michigan Survey Research Center, our partners for study recruitment and data collection.

