## Supplementary Materials for "The Effect of a U.S. Poverty Reduction Intervention on Maternal Assessments of Young Children’s Health, Nutrition, and Sleep: A Randomized Control Trial"

**Contents:**

**eMethods**

**eResults**

**eTable 1**. Marginal Effects of the Cash Gift on Dichotomous Outcomes

**eTable 2**. Descriptive Statistics and Treatment Impacts of the Poor Health Index

**eTable 3.** Factor Loadings for a Two-Factor Solution of the Poor Health Index Items

**eFigure 1.** CONSORT Diagram for Age-1, Age-2, and Age-3 Data Collection

**eMethods**

The specific amount for the cash gift was motivated by quasi-experimental work which finds a $4,000 increase in annual income (adjusted for inflation) during the first few years of life to be associated with significant boosts to children’s academic achievement^1,2^ Additionally, this amount aligns with social services many low-income families are eligible for (e.g., the Earned Income Tax Credit), highlighting the policy relevance of the study design.

At the time of recruitment, the distribution of the cash gifts was planned for the first 40 months of the children’s lives. The duration of the monthly cash gifts was subsequently extended twice. First, in response to the need to postpone in-person data collection due to the COVID-19 pandemic, the cash gifts were extended for an additional year, through 52 months. Subsequently, motivated by the evidence that the likelihood of adverse outcomes increases the more time a child experiences poverty, additional philanthropic funding extended the monthly cash transfers to 76 months.

Prior to the launch of the study, we secured approvals from state or local officials to ensure that participants would not lose eligibility for public benefits because of the cash gift, including Medicaid.^3^ A 40/60 division of the cash gifts across the sample was used to reduce study costs while also maintaining sufficient statistical power.^3^ After accounting for a predicted attrition of 20% by the final assessment (*n*=800), we are sufficiently powered to detect an effect size of 0.20 SD.^3^

***Preregistered Analyses***

**Poor Health Index.** The Poor Health Index is a pre-registered, secondary outcome available in the publicly released dataset. It is an additive index of the following items: maternal rating of the child’s overall health status on a Likert scale ranging from 1 (*excellent*) to 5 (*poor*); child disability status or diagnosis with a health condition, indicated by either 1 (*yes*) or 0 (*no*); number of doctor visits due to illness measured by either 0 (*0 – 1 visits*), 1 (*2 – 5 visits*), or 2 (*6+ visits*); number of doctor visits due to injury measured by either 0 (*0 – 1 visits*), 1 (*2 – 5 visits*), or 2 (*6+ visits*); any ER/urgent care visit in the last year indicated by either 1 (*yes*) or 0 (*no*); and the total number of ER visits measured as either 0 (*none*), 1 (*1 visit*), 2 (*2 – 5 visits*), or 3 (*6+ visits*).

**Factor Analysis.** As part of our pre-registration, we conducted an exploratory factor analysis (EFA) of the five health and healthcare items. We hypothesized the items would load onto either one “health” or two “health” and “healthcare utilization” factors. The child’s overall health rating, disability indicator, doctor visits due to illness and injury indicators, and the frequency of ER/urgent care visits were entered into an EFA with oblique oblimin rotation.

**eResults**

The coefficients below reflect the standardized treatment impact on a respective outcome, divided by the standard deviation (SD) of the control group. Marginal effects of the dichotomous outcomes derived from probit regressions are available in eTable 1.

**Diagnoses**. At each age, mothers provided information about their child’s specific health conditions and diagnoses. The most common diagnoses at Age 1 included eczema (*n*=21) and asthma (*n*=21). At Age 2, the most commonly reported diagnoses were asthma (*n*=20) and autism (*n*=9). At age 3, autism (*n*=28) and asthma (*n*=20) were again the most common diagnoses, as reported in the main manuscript. There were no statistically detectable effects of the cash gift on the likelihood of reporting a diagnosis of autism (*β*_Age3_ = 0.01, SE = 0.01, *p* = 0.76) or asthma (*β*_Age3_ = 0.01, SE = 0.01, *p* = 0.91) at Age 3.

**Vaccinations**. At each wave of data collection, mothers also responded to a dichotomous indicator of whether the child was up-to-date on routine childhood vaccinations. Between 90 – 94% of all mothers indicated their child was up-to-date on their vaccinations. There was no effect of the cash gift on maternal report of child vaccination status, β_Age1_ = -0.01, SE = 0.02, *p* = 0.67; β_Age2_ = 0.01, SE = 0.02, *p* = 0.66; β_Age3_ = 0.01, SE = 0.02, *p* = 0.51. We did not have access to medical records to confirm maternal report of vaccination status.

**Missed Medical Care.** We also surveyed the mothers on missed medical or dental care. Across time points, between 2 – 6% of all mothers reported they had missed needed medical or dental care for themselves or their child in the last year due to cost. In comparison, only 1.2% of caregivers nationally reported delaying medical care for their child due to cost (95% CI 0.9, 1.6; NCHS, 2020). There were no group differences in the likelihood of missing needed medical or dental care at any timepoint, β_Age1_ = -0.01, SE = 0.02, *p* = 0.93; β_Age2_ = 0.02, SE = 0.02, *p* = 0.40; β_Age3_ = -0.01, SE = 0.02, *p* = 0.50.

**Consumption of Cow’s Milk**. At the Age-2 visit, mothers reported on the frequency with which their child consumed unflavored cow’s milk on an average day, with options ranging from 0 (*not at all*) to 5 (*5+ times per day*). On average, mothers reported that their child consumed cow’s milk 2.11 times per day (SD = 1.62). There were no statistically detectable effects of the cash gift on the frequency of cow’s milk consumption, β_Age2_ = 0.07, SE = 0.07, *p* = 0.35

**Medicaid Receipt**. Finally, at the Age-3 visit, 71% of the sample reported receiving Medicaid. In an exploratory (non-preregistered) analysis, we detected a significantly lower Medicaid enrollment among families in the high-cash gift group. Receiving the high-cash gift resulted in a lower likelihood of being enrolled in Medicaid at Age 3, β_Age3_ = -0.07, SE = 0.03, *p* = 0.02. This effect was trending at the Age-1 (β_Age1_ = -0.05, SE = 0.03, *p* = 0.12) and Age-2 wave, β_Age2_ = -0.05, SE = 0.03, *p* = 0.08. As noted above, mothers were informed upon enrolling in the study that because the monthly cash transfers were being given as a gift, they were not taxable income and should not affect their or their child’s eligibility for Medicaid.

**Poor Health Index.** eTable 2 presents the descriptive statistics and treatment impacts of the preregistered Poor Health Index at each wave of data collection. We observed no statistically significant effect of the intervention on the Poor Health Index.

**Factor Analysis.** Neither a one- or two-factor solution appropriately fit the data. We observed poor factor loadings (eTable 3). Indeed, the KMO indicated that the variables are poorly related for factor analysis (KMO_Age1_ = 0.57, KMO_Age2_ = 0.57, KMO_Age3_ = 0.63), and the high uniqueness values suggests the variables are not well explained by the factors.

**eTable 1**. Marginal Effects of the Cash Gift on Dichotomous Outcomes

|  | | | | **Age 1** | **Age 2** | | **Age 3** | | | | | **Cumulative Impacts**  **(Age 1 – 3)** | | |
| --- | --- | --- | --- | --- | --- | --- | --- | --- | --- | --- | --- | --- | --- | --- |
| ***Health Outcomes*** | | | |  |  | |  | | | | |  | | |
|  | Diagnosed with health condition or disability | 0.03  (0.02) | | | | -0.01  (0.02) | | 0.03  (0.02) | | | | | 0.02  (0.03) | |
| ***Healthcare Utilization*** | | | | | | | | | | |  | | | |
|  | 2+ doctor visits due to illness | | 0.05  (0.03) | | -0.01  (0.03) | | | | -0.01  (0.03) | | | | | 0.01  (0.02) |
|  | 2+ doctor visits due to injury | | 0.01  (0.01) | | -0.01  (0.01) | | | | -0.02 (0.02) | | | | | -0.01  (0.01) |
| ***Non-Preregistered Outcomes*** | | |  | |  | | | |  | | | | |  |
|  | Up-to-Date on Vaccinations | | -0.01  (0.02) | | 0.01  (0.02) | | | | 0.01  (0.02) | | | | | 0.01  (0.01) |
|  | Missed Medical/Dental Care | | 0.01  (0.02) | | 0.02  (0.02) | | | | -0.02  (0.02) | | | | | 0.01  (0.01) |
|  | Medicaid Enrollment | | -0.05  (0.03) | | -0.05+  (0.03) | | | | -0.07*  (0.03) | | | | | -0.06*  (0.02) |
|  | Autism Diagnosis | | N/A | | N/A | | | | 0.01  (0.01) | | | | | N/A |
|  | Asthma Diagnosis | | N/A | | N/A | | | | 0.01  (0.01) | | | | | N/A |
| **Sample Size** | | | | **929** | **919** | | **920** | | | **2,768** | | | | |

*Note*. + *p*<0.10, **p*<0.05. Dichotomous outcomes were estimated using logistic regression and the coefficients represent marginal effects. Standard errors are reported in parentheses. Estimates are adjusted for the covariates listed in Table 1, site-based fixed effects, survey administration method (i.e., phone or in-person) at the Age 1 survey, and child age at the time of the assessment.

**eTable 2**. Descriptive Statistics and Treatment Impacts of the Poor Health Index

|  | **Mean (SD)** | | **Treatment Impacts** |
| --- | --- | --- | --- |
|  | *Low-Cash Gift* | *High-Cash Gift* | ES (SE) |
| Age 1 | 5.56  (2.03) | 5.75  (2.22) | 0.12  (0.07) |
| Age 2 | 4.91  (1.91) | 4.93  (1.98) | 0.02  (0.14) |
| Age 3 | 4.92  (2.12) | 4.92  (1.96) | 0.04  (0.07) |
| **N** | **542 – 547** | **372 – 382** | **919 – 929** |
| *Note.* +*p*<0.10, **p*<0.05. The composite is an additive index of the following items: child’s health rating ranging from 1 (*excellent*) to 5 (*poor*); child disability status or diagnosis with a health condition, 1 (*yes*) or 0 (*no*); number of doctor visits due to illness, 0 (*0 – 1 visit*), 1 (*2 – 5 visits*), or 2 (*6+ visits*); number of doctor visits due to injury, 0 (*0 – 1 visit*), 1 (*2 – 5 visits*), or 2 (*6+ visits*); any ER/urgent care visit in the last year 1 (*yes*) or 0 (*no*); and the total number of ER visits, 0 (*0 visits*), 1 (*1 visit*), 2 (*2 – 5 visits*), or 3 (*6+ visits*). ES reflects the standardized difference between the two groups, divided by the SD of the control group. Robust standard errors are in parentheses. Estimates are adjusted for the covariates listed in Table 1, site-based fixed effects, survey administration method (i.e., phone or in-person) at the Age 1 survey, and child age at the time of the assessment. | | | |

**eTable 3.** Factor Loadings for a Two-Factor Solution of the Poor Health Index Items

|  | **Age 1** | | | **Age 2** | | | **Age 3** | | |
| --- | --- | --- | --- | --- | --- | --- | --- | --- | --- |
|  | *Factor 1* | *Factor 2* | *Uniqueness* | *Factor 1* | *Factor 2* | *Uniqueness* | *Factor 1* | *Factor 2* | *Uniqueness* |
| Child Health Rating | -0.06 | 0.43 | 0.84 | -0.10 | 0.41 | 0.86 | -0.07 | 0.46 | 0.81 |
| Disability Diagnosis | -0.01 | 0.33 | 0.89 | -0.01 | 0.37 | 0.86 | -0.05 | 0.40 | 0.86 |
| 2+ doctor visits due to illness | 0.19 | 0.44 | 0.69 | 0.14 | 0.47 | 0.69 | 0.24 | 0.42 | 0.65 |
| 2+ doctor visits due to injury | -0.07 | 0.15 | 0.97 | 0.01 | 0.32 | 0.89 | 0.11 | 0.23 | 0.91 |
| Any ER/Urgent Care Visit | 0.94 | -0.08 | 0.17 | 0.95 | -0.07 | 0.15 | 0.95 | -0.06 | 0.15 |
| Number of ER/Urgent Care Visits | 0.87 | 0.12 | 0.13 | 0.89 | 0.09 | 0.12 | 0.91 | 0.07 | 0.11 |
| Eigenvalue | 1.94 | 0.35 |  | 1.97 | 0.44 |  | 2.10 | 0.40 |  |

*Note*. +*p*<0.10, **p*<0.05. Exploratory factor analysis was conducted with oblique oblimin rotation.

**eFigure 1.** CONSORT Diagram for Age-1, Age-2, and Age-3 Data Collection

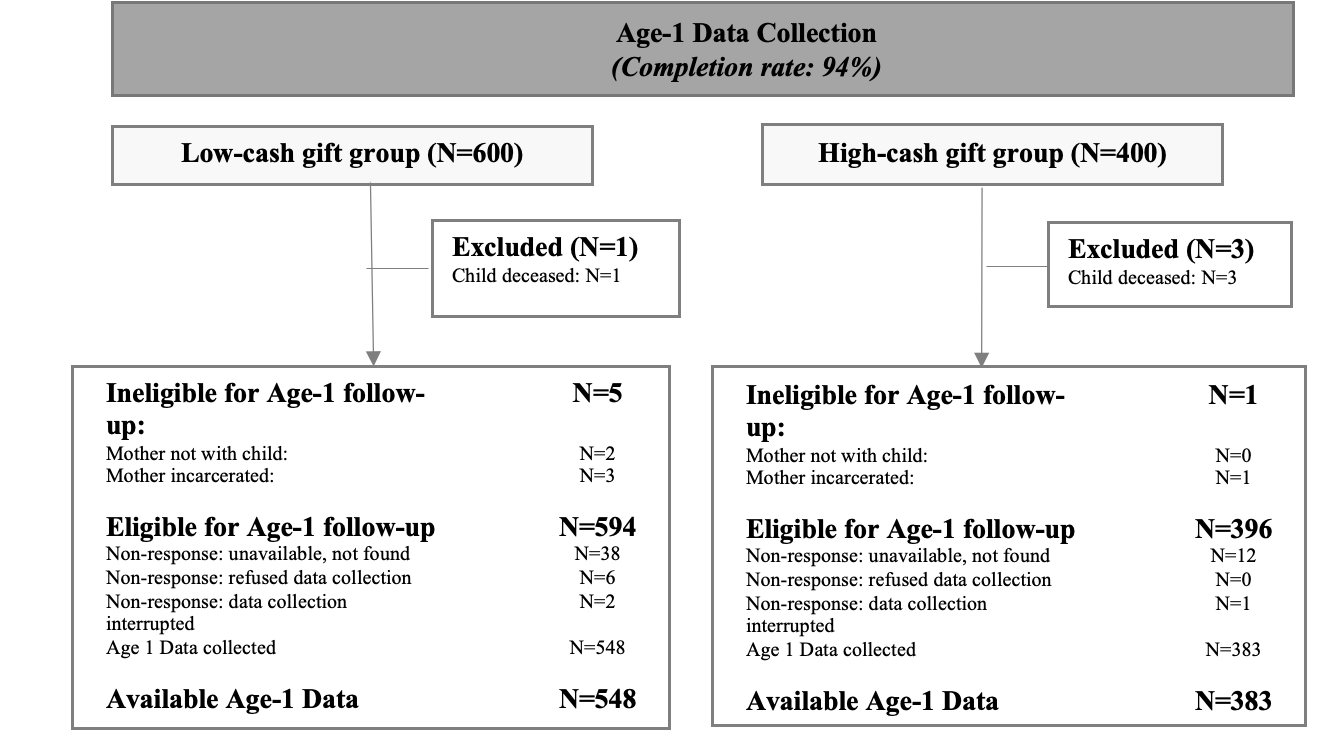

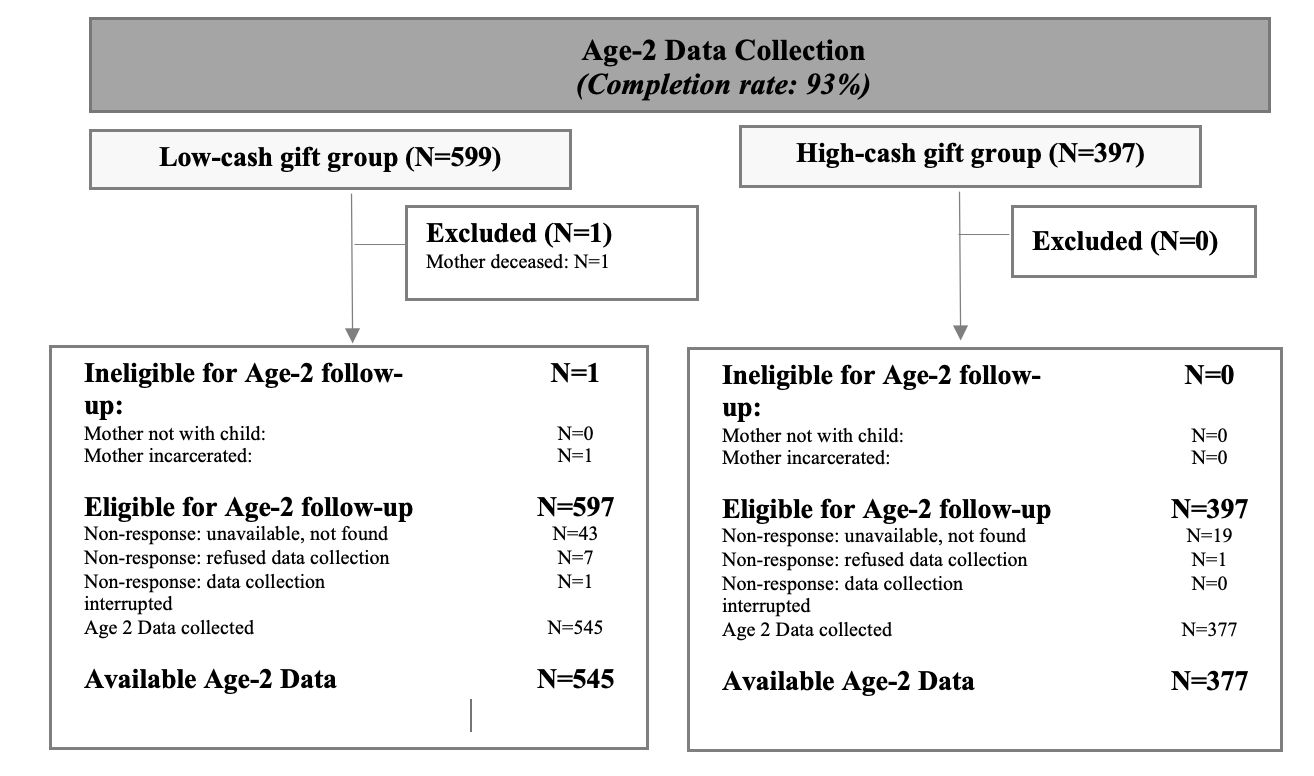

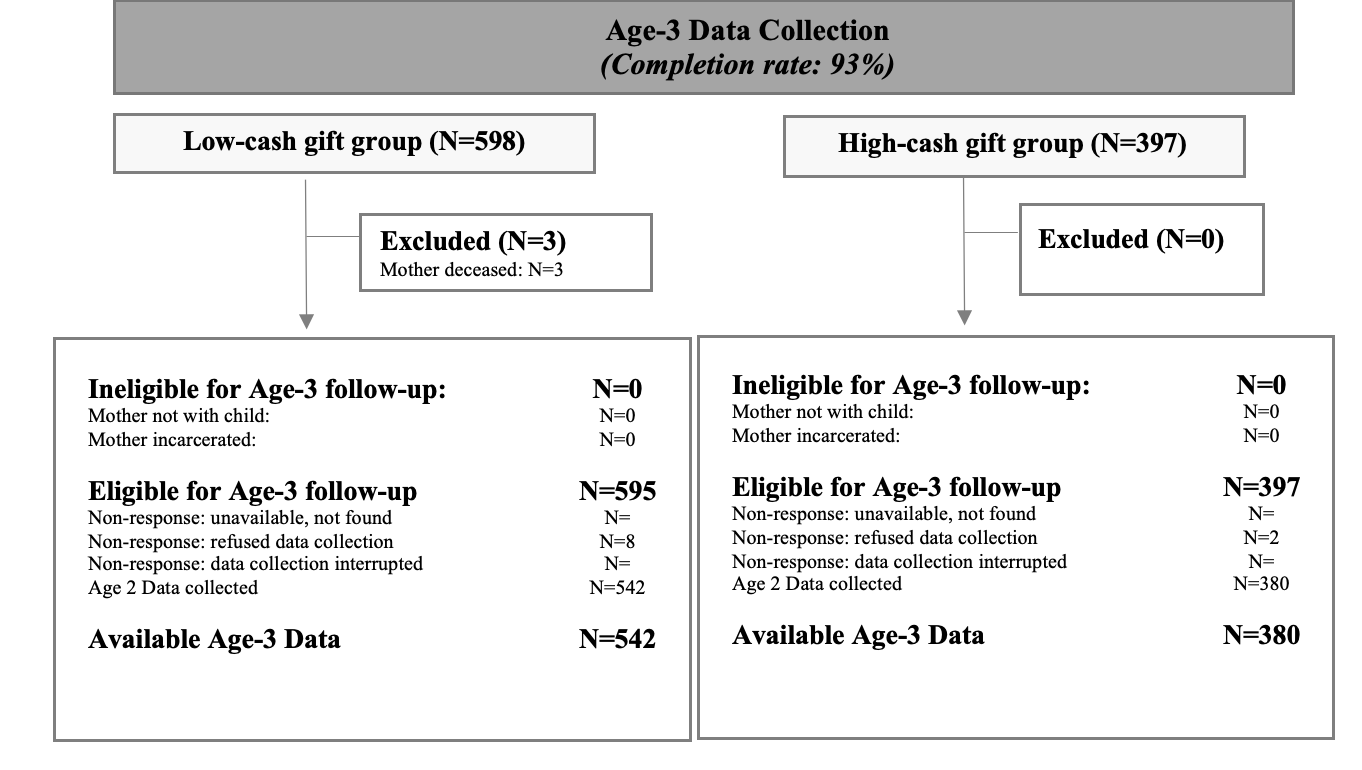
